# Cohorting of Non-Critically Ill COVID-19 Patients: A Multicenter Survey Study (COVID-COHORT)

**DOI:** 10.1101/2020.11.01.20194233

**Authors:** Ushma Purohit, Mike Fralick

## Abstract

The COVID-19 pandemic has posed novel infection-control challenges for hospitals around the globe. One infection-control strategy that has been widely used in the context of other outbreaks is patient cohorting. This strategy refers to the placement of all patients exposed to the same laboratory-confirmed infectious agent in one location within the hospital. Little is known about the current utilization of this strategy with non-critically ill COVID-19 patients. An international multicenter, survey study was conducted to identify what strategies are planned or in place for patients with COVID-19 who are not critically ill. The survey was distributed from March 23-29th, 2020 to GIM physicians in Canada, USA, Denmark, Singapore, Hong Kong, and England. Of the 31 hospitals, 29 (94%) indicated that they plan on cohorting all GIM patients with COVID-19 to one location in the hospital. Among these 29 hospitals, 23 (79%) had implemented the plan at the time of the survey. The primary reasons for this decision were to limit the spread of COVID-19 and conserve PPE use. In conclusion, in the face of a novel virus there is near unanimity in the practice of patient cohorting as a potential mitigation strategy.

## Introduction

Severe acute respiratory syndrome coronavirus 2 (SARS-CoV-2) is a novel coronavirus that has been identified as the cause of the coronavirus disease 2019 (COVID-19). As of May 24, 2020, there were over 5.31 million cases reported worldwide and over 342,000 deaths.^1^

Patient cohorting is an infection control strategy frequently employed in the context of the current COVID-19 outbreak. There are no formal guidelines on whether patients who are non-critically ill should be cohorted in the same geographic location. However, The Centre for Disease Control and the European Centre for Disease Prevention has advised that facilities consider designating units within the hospital, along with dedicated healthcare providers (HCP), to care for known or suspected COVID-19 patients.^2^ This recommendation has been made to limit HCP exposure and to conserve Personal Protective Equipment (PPE).^2^ The objective of our study was to determine the current policies and procedures enacted for cohorting of non-critically ill patients.

## Methods

We conducted an international, multicenter, survey study to identify the strategies planned (or in place) for patients with COVID-19 who are not critically ill and admitted to General Internal Medicine (GIM). We focused on GIM because this is the most common service caring for non-critically ill patients with COVID-19 based on data from countries that were affected earlier by COVID-19.

The survey was distributed from March 23-29^th^, 2020 via email. Survey respondents included GIM physicians from Canada, USA, Denmark, Singapore, Hong Kong, and England. Physicians were not asked for any self-identifying information and were ensured that results would be reported at the regional level and not at the hospital-level. The survey included multiple-choice questions (Yes/No) about whether the physician’s hospital had plans for patient cohorting and whether the plans had already been implemented. They were also asked to provide reasoning behind their hospital’s decision on whether to implement patient cohorting using an open-ended short-answer format. The study was exempt from Research Ethics Board approval. Descriptive statistics were used to summarize the data.

## Results

Responses were provided from 31 hospitals, of which 20 were from Canada (15 from Ontario, Canada). Of the 31 hospitals, 29 (94%) indicated that they plan on cohorting all GIM patients with COVID-19 to one location in the hospital. Among these 29 hospitals, 23 (79%) had implemented the plan at the time of the survey. The main reasons for cohorting were to minimize nosocomial spread of COVID-19 and conserve PPE. Six hospitals did not have any plans to implement patient cohorting. Survey respondents stated this was because there were no admitted individuals with COVID-19 at the time of completing this survey.

## Discussion

In the face of a novel virus there is near unanimity in the practice of patient cohorting as a potential mitigation strategy. Of the 31 included hospitals, 29 have decided to cohort patients onto one geographic location. The primary reasons for this decision were to limit the spread of COVID-19 and conserve PPE use.

Our findings align with the measures taken by several hospital sites during the Taiwan SARS outbreak.^3^ One study reported 0.03 cases/bed in hospitals with cohorting vs 0.13 cases/bed in hospitals without (P<0.03).^4^ Patient cohorting, by virtue of permitting PPE resuse, has also been shown to decrease PPE shortages. According to the World Health Organization, in the event of PPE shortages, HCPs may wear the same respirator while caring for multiple patients with COVID-19, in the same isolated location.^5^ Cohorting may also contribute to containing the widespread contamination of the healthcare environment that has been documented in the setting of SARS-CoV-2.^6^ Specifically, in areas of the hospital where patient cohorting occurred, the ability to detect SARS-CoV-2 was 4-fold higher than in areas where there were no patients with COVID-19.^6^ Important limitations to our study include the lack of a systematic survey strategy and the lack of information on the rate of nosocomial outbreaks of respiratory pathogens within the participating hospitals.

As COVID-19 continues to pose novel challenges for hospitals across the globe, it becomes vital for healthcare communities to share best practices and learn from one another. Our multicenter, international survey identified that all hospitals with active cases of COVID-19 on GIM wards are practicing patient cohorting.

## Data Availability

N/A

## Acknowledgements

We would like to thank Dr. Christopher Kandel, Dr. Michael Colacci, Dr. Sagar Rohailla and Nancy Figueroa for their feedback on earlier versions of the manuscript.

